# Perceptions of frailty and frailty screening of people living with HIV and their healthcare professionals in Ethiopia: A qualitative study

**DOI:** 10.1101/2025.07.08.25331102

**Authors:** Clea Kapos Tanner, Kate Alford, Endrias Markos Woldesemayat, Netsanet Abera Asseffa, Ephrem Lejore, Stephanie Marquart, Natalie St. Clair-Sullivan, Andargachew Kassa, Taye Gari, Kenene Gutema, Nana Chea, Jaime H. Vera

**Affiliations:** University of Sussex, BSMS, Brighton, United Kingdom; Hawassa University, School of Public Health, Hawassa, Ethiopia; Hawassa University, School of Midwifery Nursing, Hawassa, Ethiopia

**Keywords:** Ageing with HIV, HIV and frailty, southern Ethiopia, perceptions of frailty, frailty screening methods

## Abstract

**Background:** People with HIV (PWH) are an ageing population with increased risk of comorbidities, like frailty. By 2040, 25% of PLWH in sub-Saharan Africa will be over 50 years old, posing new challenges for healthcare systems. This study aims to explore perceptions of frailty and frailty screening among PWH and healthcare professionals (HCPs) in southern Ethiopia.

**Methods:** In-depth interviews and focus groups were conducted with PWH and HCPs recruited from three hospitals in southern Ethiopia. Pseudonymised transcripts were analysed using reflexive thematic analysis.

**Results:** 26 PWH and 12 HCPs participated in interviews or focus groups. Both groups noted that frailty is not discussed in HIV settings. HCPs feared discouraging clients by discussing frailty, though both groups favoured increased solution-focussed discussions. PWH cited socio-economic challenges as key influencing factors and impacts of frailty.

**Conclusions:** The findings underscore the need for increased frailty discussions and holistic frailty interventions integrating physical and social support.

## Introduction

The global development and uptake of combination antiretroviral therapy (cART) has transformed the prognosis and life expectancy of many people with HIV, leading to an increased number of older people with HIV. ^1^ People with HIV experience age-related co-morbidities and geriatric syndromes, such as mobility issues, polypharmacy, decreased cognitive function and frailty, at disproportionately higher rates and at younger ages than the general population. ^2–5,11^ Two thirds of the global population of people with HIV live in sub-Saharan Africa,^6,7^ including an estimated 610,000 people in Ethiopia.^8^ It is projected that by 2040, older people with HIV, classified as people aged 50 and over by UNAIDS, will make up 25% of those living with HIV in this region.^7–10^ Along with physical challenges associated with age-associated syndromes, stigma associated with both HIV and ageing, combined with the potential for age-related discrimination, augments challenges for care provision and promoting healthy ageing in this population.^4,11^

Traditionally, in Ethiopia, care of older people would be the responsibility of their extended family unit. However, this community is affected by an increasing ageing population and limitations on younger family members’ ability to care for their older relatives.^12,13^ This shift in care for older people with HIV increases strain on healthcare systems and requires further understanding to improve the provision of geriatric care in this cohort. Consequently, aging people with HIV represent a critical population for advancing our understanding of care needs in Ethiopia. The development and adaption of care systems to address these evolving needs are further hindered by the limited healthcare resources available throughout Ethiopia and by the pervasive stigma associated with HIV.^14,15^

Frailty has been identified as a marker of vulnerability in people with HIV, particularly older people with HIV, and as a priority focus for clinical care of this population.^16^ Frailty is characterised by declines in multiple physiological systems and reserves, which decrease the individual’s ability to compensate for stressors, such as falls, that cause deterioration in health and function.^17^ Frailty has also been shown to associated with decreased health-related quality of life for people with HIV in other countries.^18^ This association with lower quality of life could be linked to frailty-related pain, limitations on lifestyle factors including work and social life, and financial disadvantages.^18,19^

Prevalence of frailty in a cohort of 187 people with HIV in southern Ethiopia has been reported to be 9.1%. However, over two thirds (75.4%) of the cohort were identified as pre-frail^20^, indicating a high burden of frailty in this region. Aside from the aforementioned study, the vast majority of frailty research has been conducted in the global North and we found no research pertaining to perceptions of frailty in Ethiopia.^20^ Other studies have investigated the prevalence of and factors influencing frailty in other areas of sub-Saharan Africa.^21^ Indeed, a qualitative study in Zambia explored the social construct of ageing for people with and without HIV and found that frailty was considered to be an “automatic progression” from the impact of the HIV infection. Furthermore, the report found that some individuals without HIV avoid taking medication for frailty out of concern that in doing so might be perceived as taking ART.^22^

Understanding the cultural nuances surrounding age and frailty, from both community and healthcare perspectives in Ethiopia, will facilitate conception of culturally sensitive and effective frailty interventions to support patient wellbeing and improve quality of life. This exploratory study aims to investigate perceptions of frailty among people with HIV, assess whether frailty is currently addressed in healthcare, explore the attitudes of both people with HIV and HCPs toward discussing frailty, and examine frailty screening method preferences.

## Methods

### Study Design

We conducted an exploratory qualitative study with participants recruited from three HIV clinics in southern Ethiopia in November 2021. People with HIV and their HCPs took part in either in-depth interviews or focus groups.

### Participants

Participants were people with HIV attending specialist HIV clinics, aged 35 and over and on cART. The HIV HCPs were working at one of the three recruitment sites.

### Study Procedure

Study participants were identified by the HIV clinical teams at one of three participating hospitals in the Sidama region of Ethiopia: Hawassa University Comprehensive Specialized Hospital, Yirgalem Hospital, and Adare Hospital, and then referred to the research team. All participants gave informed consent before taking part in either an in-depth interview or a focus group. All interviews and focus groups were conducted in-person and audio-recorded in Amharic, then transcribed into English. To help structure the interviews, the research team developed interview guides, which included open-ended questions, as well as specific probes. The key areas of investigation were understanding of frailty, impacts of frailty and perceptions of discussing frailty and frailty screening (See table 1 for the interview guide, including questions and probes).

**TABLE 1.** Interview Guide – Key Questions and Probes.

|  |
| --- |
| **1. How would you describe frailty?**<br>- What comes to your mind when you hear frailty?<br>- What does frailty mean to you? What do you know about it?<br>- What does it let you think?<br>**2. How would you describe your feelings about frailty?**<br>- Does it make you feel good or bad, why?<br>- Could you tell me if there is a change in your way of life or practice that changed due to these feelings?<br>**3. In your opinion, what factors do you think influence frailty?**<br>(Ask one by one)<br>- Internal factors such as Age, Sex, Employment status, Duration of HIV illness, Type of medical care received<br>- External factors<br>- Environmental factors<br>- Weather condition<br>- Malnutrition/famine<br>- Displacement or security problems<br>- Support networks<br>- Stigmatization<br>**4. How problematic is frailty in your opinion compared to your other major concerns?**<br>- Is frailty a concern for you in the future? If so, could you explain your concern further?<br>- In your opinion, do you think that a person with HIV will change their lifestyle due to frailty?<br>**5. In your opinion does frailty is related to anything?**<br>- Do you think there is an association between frailty and HIV treatment status? If so, can you describe it?<br>**6. Have you ever discussed frailty with anyone before?**<br>- If so, when did you discuss it? Do you think there is right time to discuss it?<br>- How did your discussion make you feel?<br>- Was your conversation good or bad?<br>- If you have never discussed it before, how does it make you feel to discuss it?<br>**7. Do you think health care providers should start talking to people on HIV treatment about frailty?** If so, do you think there is a right age group or reason to talk? |

### Data Analysis

The study team conducted analysis of the interview and focus group transcripts in order to identify themes relating to the perceptions of frailty in people with HIV in Ethiopia. Transcripts from both interviews and focus groups were analysed concurrently, using the same coding frame, to allow for full exploration of the similarities and differences in the views expressed in each context. Thematic Analysis, informed by Braun and Clark’s six phase process^23^ was conducted by SM and KA. Phase 1 involved familiarisation with the dataset, reading and rereading for an immersive approach. Phase 2 involved coding (generating succinct labels, or codes, that represent principal features of the dataset). Phase 3 consisted of building initial themes by examining the codes and identifying broader patterns among them as potential themes. Phase 4 required continued development and review of the themes. During this iterative process of reviewing the coding and themes, which required going back and forth between different phases, the themes were checked against coded transcripts, redefined, further developed, combined, split or abandoned. The final themes were defined and named in Phase 5, with detailed analysis of each theme, including their scope, focus and specific wording. The final themes were decided by KA and CT, who examined similarities and differences of codes within themes independently to define final theme names and content. The last phase involved writing up by creating the analytic narrative and selecting data extracts along with contextualising the data in other relevant research. To limit loss of cultural and linguistic nuance, those who conducted the interviews and focus groups were routinely questioned by those conducting the analysis regarding the codes identified and where any ambiguity in the data was felt. Lastly, a session was further organised with these team members to finalise themes and ensure that the results presented reflected the impressions they gained during the interview process.

## Results

A total of 38 participants were recruited. 14 people with HIV and 5 HCPs took part in in-depth interviews, which lasted between 20-58 minutes. In addition, 12 people with HIV and 7 HCPs took part in three focus groups: i) 6 male people with HIV, ii) 6 female people with HIV, iii) 7 mixed HCPs. The focus groups lasted between 55-66 minutes. The people with HIV included 11 female patients and 14 male patients (and one participant with no gender data), with an age range of 36-65. The HCP group included 3 male HCPs and 9 female HCPs with job roles including medical doctor, nurse, infectious diseases specialist and laboratory worker (please see table 2 for full participant demographics).

**TABLE 2.** Participant Characteristics.

| Characteristics | n (%) – unless noted otherwise |
| --- | --- |
| *People with HIV (PWH) Participants (n=26)* |  |
| Age (years) [median (range)] | 54.5 (36-65) |
| Gender |  |
| Male | 14 (53.85) |
| Female | 11 (42.3) |
| No Data | 1 (3.85) |
| Years on cART (range) | 5-24 |
| *Healthcare Professionals (n=12)* |  |
| Age (years) [median (range)] | 40 (30-50) |
| Gender |  |
| Male | 3 (25) |
| Female | 9 (75) |
| Job Role |  |
| Nurse | 9 (75) |
| Infectious Disease Specialist | 1 (8.33) |
| Medical Doctor | 1 (8.33) |
| Lab Worker | 1 (8.33) |
| Duration in profession<br>(years)[median(range)] | 5 (1-17) |

Three main themes were identified with a view to understanding people with HIV and HCP’s perceptions of frailty and addressing frailty in healthcare settings. These themes are summarised in the following sections (please see table 3 for participant quotations supporting the themes).

**TABLE 3.**
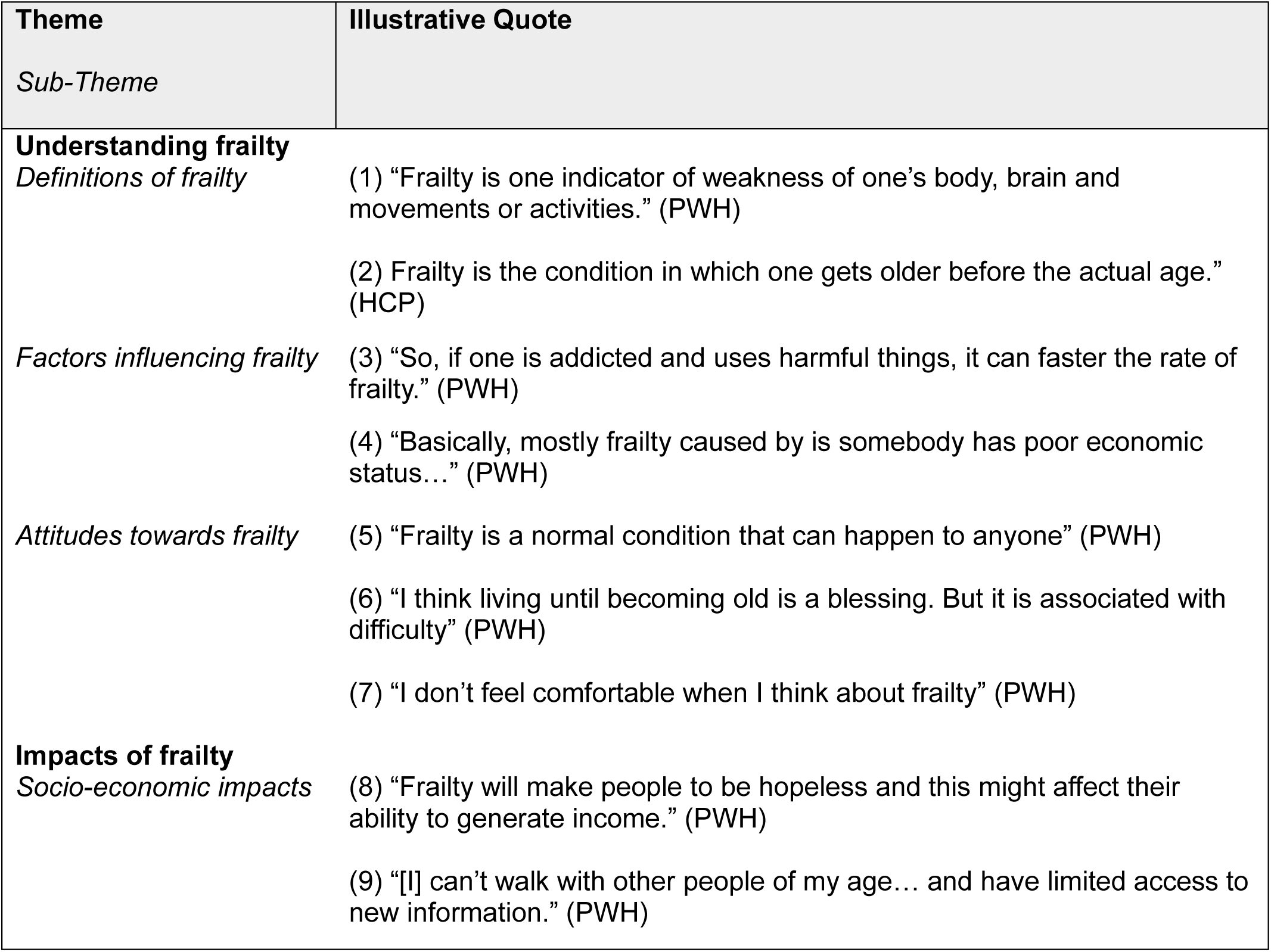
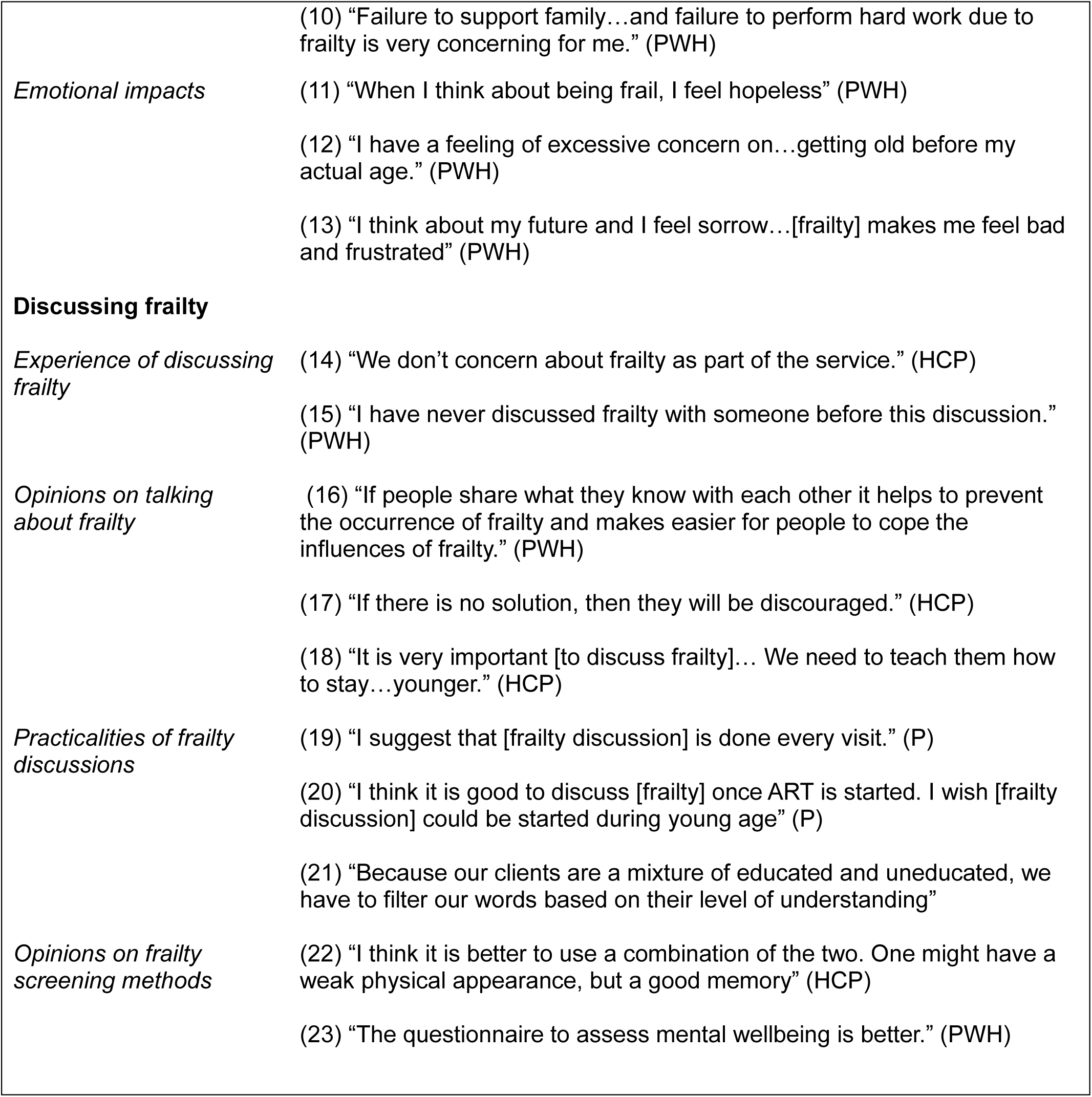
Themes with illustrative quotes.

## Theme 1: Understanding Frailty

### Definitions of frailty

There was some variation across the participants’ understanding of the term ‘frailty’. Frailty was described as a state of mental and physical deterioration, associated with weakness and physical discomfort. There was a contrast between most people with HIV who described frailty as premature ageing, while others believed it to be an unavoidable consequence of ageing. Whereas HCPs had a more homogenous understanding of frailty; they also described frailty as premature ageing and mentioned limitations in completing day-to-day tasks as a defining feature.

### Factors influencing frailty

Regarding factors that influence frailty, there was distinction between those who believe it is an unavoidable result of ageing and those who thought it is influenced by various social, economic or medical factors. People with HIV and HCPs both mentioned poverty, employment status and financial support as economic influences on frailty development. Social factors raised by both groups included stigma, living conditions, displacement, social networks and family caring responsibilities. Though generally people with HIV thought that displacement and insecurity have no impact on frailty. When considering medical factors, in addition to general poor health, malnutrition and alcohol and drug misuse were identified as factors influencing frailty by people with HIV. HCPs also suggested female patients were more likely to experience frailty. Participants in both groups also considered stress to be a factor affecting frailty. Time living with HIV was also identified as a factor influencing frailty by people with HIV and HCPs, but when asked directly about links between HIV and frailty, many people with HIV considered the two to be unrelated.

People with HIV consistently mentioned God when discussing frailty. Many participants felt that they did not have control over their propensity to develop frailty and felt God determines how long they will live and at what point they will age. This view generally coincided with the definition of frailty as an unavoidable consequence of ageing.

### Attitudes towards frailty

There was a further contrast in attitudes towards frailty. While most participants held a more negative attitude towards frailty, describing feeling a sense of hopelessness when they think about frailty as an indicator of premature old age, one participant discussed frailty as a positive indicator of achievement in surviving to an older age. Interestingly, while some participants were not directly concerned about frailty, when directly prompted to talk about feelings around frailty, all participants linked frailty with general negative emotion.

## Theme 2: The Impacts of Frailty

### Socio-economic impacts

Several people with HIV were focussed on the financial impact of frailty. They cited lack of employment, and therefore lack of income, resulting from physical and mental inability to complete tasks, as a significant financial impact of frailty. When asked about potential lifestyle changes due to frailty, many people with HIV thought that frailty would not impact lifestyle at all. However, lack of employment was described as one potential lifestyle change by some. Discussions around income and employment status also raised concerns about being able to support a family and general economic security due to frailty. In addition to economic impacts, some people with HIV also mentioned isolation from their peers as a result of any changes in physical and mental capabilities as a potential impact. This also led to discussion about not having access to information because of isolation from the wider community.

### Emotional impacts

Several people with HIV also discussed how mental wellbeing may be affected by frailty, with feelings such as hopelessness, loneliness and fear being associated with frailty. Increased stress was also identified both as a potential exacerbator or impact of frailty. Several people with HIV discussed further consequences from emotional impacts of frailty, such as hopelessness leading to lower income or stress impacting an individual’s capacity to work.

## Theme 3: Discussing Frailty

There were detailed questions about discussing frailty: how participants felt about it and the practicalities of how discussions about it should be conducted.

### Experience of discussing frailty

Participants across both groups commented that frailty is not currently spoken about either in healthcare settings or within community or family settings. HCPs shared they don’t currently discuss frailty with patients, instead asking about general health concerns.

### Opinions on talking about frailty

People with HIV expressed positive feelings about discussing frailty more. They felt that discussing frailty should not be difficult and would be helpful for delaying frailty, knowledge sharing and potential solutions. HCPs also felt there were lots of potential benefits to general frailty discussions, including information sharing, potential for prevention and relief, and helping people with frailty feel less alone. However, most HCPs further expressed concern that patients do not want to discuss frailty and fear offending patients by discussing frailty directly. The main priority for them was to ensure that discussions about frailty include potential solutions, as they expressed concern that direct discussion without solutions could negatively affect patients’ morale. Accordingly, they also identified adjusting their language use for each patient’s understanding and avoiding jargon, as priorities for keeping frailty discussions accessible and positive for their patients. Although all patient-facing HCPs felt confident in discussing frailty with patients who are comfortable with the topic, they also raised education for HCPs as a tool for increasing HCP confidence and thereby improving discussion about frailty.

### Practicalities of frailty discussions

When asked about the practicalities of frailty discussions in healthcare settings, HCPs and people with HIV thought that frailty should be discussed regularly, with people with HIV suggesting timeframes from annually to as frequently as monthly. People with HIV also proposed a wide range of durations for discussions about frailty, with some suggesting that longer than five minutes of discussion could make frailty seem like a larger issue than they felt it was, whereas others recommended that discussions should last up to an hour to allow enough time for a full discussion and questions. Both participant groups also suggested a variety of ages that they deemed suitable for initial discussions about frailty. These ranged from 20 years old to over 50, but HCPs generally specified over 25 as an appropriate age to begin discussing frailty with their HIV patients. HCPs also identified other potential trigger points for discussing frailty beyond age. They suggested introducing frailty discussions at the point of HIV diagnosis, when patients are collecting medication, and if frailty becomes relevant to the patient’s specific care plan. People with HIV were also asked which type of healthcare professional they would prefer to discuss frailty with. Responses were varied, but included doctors, HCPs with specific HIV knowledge, a multidisciplinary team of HCPs and HCPs with HIV themselves.

### Opinions on frailty screening methods

When asked for their opinions on frailty screening methods, including mental wellbeing assessments and physical assessments, all HCPs advocated for using both screening methods. They explained that patients may show signs of frailty in either mental wellbeing or physical assessments, but not necessarily both, and therefore using both methods would be more effective for detecting all signs of frailty. People with HIV were more divided with some favouring the physical assessment and others preferring the mental wellbeing assessment, with only one participant suggesting using both assessment methods. Some participants felt that physical tests can be annoying, whilst others felt that mental wellbeing tests could be offensive. Interestingly, those in individual interviews tended to prefer mental wellbeing assessments, whereas the participants of both focus groups suggested the physical wellbeing assessment would be more useful.

## Discussion

This study found that people with HIV in southern Ethiopia would welcome more discussion about frailty and perceived increased discussion about frailty as beneficial for potential prevention and management. Healthcare professionals also felt more discussion about frailty would be positive for increased awareness of frailty, however they were concerned that these conversations should be solutions-focussed to avoid discouraging their patients. Socio-economic impacts of frailty, such as impacts on income and isolation, were the main concerns about frailty for people with HIV and similarly socio-economic factors, such as nutrition and employment status, were identified as the key influences on frailty. Both HCPs and individuals with HIV supported increased frailty screening. HCPs strongly favoured the use of both mental and physical wellbeing assessments. Whereas individuals with HIV expressed a preference for either mental or physical assessments, with no clear consensus for one over the other across the cohort.

This study’s finding that individuals with HIV and their HCPs support increased discussion of frailty, particularly with a solutions-focussed approach, aligns with broader research on perceptions of frailty. Multiple recent studies in the UK highlight that people with HIV would welcome more information about frailty and particularly management of frailty.^24–26^ Similarly a study in Zambia on the construct of ageing posits increased communication about age-related issues to reduce ageism and improve health outcomes for older people with HIV.^27^ While there is consistency in receptiveness to frailty discussion, views on the influences on frailty are more varied. This study found people considered socio-economic factors, such as nutrition, substance misuse and employment status, to be the greatest factors that influence frailty. In contrast, the study in Zambia found that frailty was perceived as an ‘automatic’ result of long-term HIV.^27^ While some HCPs and people with HIV in the present study identified time living with HIV as a factor influencing frailty, the majority participants did not feel there was a link between HIV and frailty, beyond adherence to cART, when asked about the association directly. This contrast between the two studies highlights that frailty is a unique concept in different settings and therefore solutions for frailty must also be tailored for each unique setting.

Overall, people with HIV wanted more information about frailty and more opportunities to discuss frailty to aid in its prevention and management. It is important to consider that their major concerns about frailty centred around socio-economic impacts of and influences on frailty. The most common worries were maintaining sufficient income to support themselves and their families and avoiding isolation. This suggests a greater focus on socio-economic wellbeing in this region and aligns with research that points out lack of social care systems and economic support which may exacerbate the challenges faced by ageing individuals this region.^15,28^ As individuals in Ethiopia move away from traditional extended family care and the lack of formal systems to address age-related issues, like frailty, older individuals are left increasingly vulnerable and isolated from their communities.^15^ Therefore, frailty management in this region must incorporate multifaceted solutions that include social support as a crucial element, in addition to any physical interventions. There is movement towards frailty being considered more holistically in HIV settings, with calls for psychological factors to be considered equally with the physical.^26,29^ This study demonstrates that future frailty interventions for this region must go further and incorporate broader socio-economic concerns to prioritise independence and daily function for ageing people with HIV.

This study found that, while some HCPs reported discussing frailty informally with patients, frailty is not currently formally discussed in HIV healthcare settings in southern Ethiopia. This is likely due to the higher prevalence of frailty at younger ages among people with HIV than the general population, meaning that HCPs are less likely to raise age-related issues, including frailty, with these younger patients.^20^ Furthermore, HCPs discussed their concern about offending or discouraging patients by raising frailty with them. However, the range of ages suggested by patients, starting as low as 20, for initiating discussions about frailty, suggests that people with HIV are more comfortable discussing frailty at younger ages than HCPs assume. Healthcare workers generally stated that they felt confident in discussing frailty, but the concern about offending or discouraging patients could indicate a lack of confidence in offering advice about effective frailty management. This has been the case in other settings where HIV professionals felt they were not best placed to provide frailty care as they were not experts in geriatric care.^24^ Therefore, HCP’s confidence in leading solutions-focussed discussions about frailty could benefit from further training. This training should increase HCPs awareness of their patients’ reported comfort with discussing frailty and focus on applying effective frailty interventions with a patient-centred approach.

This study benefits from a diverse sample of people accessing HIV services in this region of Ethiopia. However, the analysis relied on translated interviews and focus groups, which may have limited the comprehension of potential linguistic and cultural nuance. The research team attempted to mitigate this, by consulting with the interview team in Ethiopia at different stages of the analysis, to ensure the results presented were consistent with their understanding during the interview process. Furthermore, as participants were recruited from clinics in one region of Ethiopia, these results are not more widely generalisable across this large and culturally diverse country.

Frailty is not currently discussed formally in HIV healthcare settings in this region of Ethiopia and people with HIV would generally welcome more open discussion about frailty to aid knowledge sharing about prevention and management of frailty. HCPs also expressed interest in increased awareness about frailty but highlighted the need for frailty discussions to be solutions-focussed to avoid discouraging patients. This study highlights the socio-economic challenges associated with frailty in this region and demonstrates the need for holistic frailty interventions. Effective frailty solutions should not only focus on physical health but also integrate broader social support to address the multifaceted nature of the issue. A more comprehensive approach to frailty prevention and management would support individuals with HIV to maintain their independence and develop strategies that facilitate daily functioning.

## Data Availability

Data in the present study are available upon request in the form of anonymized transcripts a.

